# Genetic proxies of GLP1R expression in whole blood are associated with lower risk of psoriasis and psoriatic arthritis

**DOI:** 10.1101/2025.10.12.25337838

**Authors:** Ravi Ramessur, Alya GA Arham, Jake Saklatvala, Laurent Thomas, Ben M Brumpton, Mari Hoff, Vibeke Videm, Bjørn Olav Åsvold, Daniel Shin, Dokyoon Kim, Michael A Simpson, Catherine H Smith, Mari Løset, Joel M Gelfand

## Abstract

Psoriasis and psoriatic arthritis (PsA) are chronic immune-mediated inflammatory diseases associated with obesity, cardiometabolic disease, and mortality. The glucagon-like peptide-1 (GLP1) pathway has emerged as a key factor in the development of obesity and cardiometabolic disease. GLP1 receptor agonists demonstrably improve insulin resistance, dyslipidemia, obesity, and mortality, with emerging evidence suggesting potential benefit for psoriasis. To evaluate whether GLP1 biology influences psoriatic disease risk, we conducted a Mendelian randomization study.

A 22-variant cis-eQTL genetic instrument for GLP1R expression in whole blood from the eQTLGen consortium was generated as a proxy for GLP1R pathway activity (n=31,684). Genetic proxies of GLP1R expression were associated with a lower risk of psoriasis (OR=0.723, 95% CI 0.678-0.771, p=1.08×10^−22^) and PsA (OR=0.483, 95% CI 0.402-0.580, p=5.33×10^−15^) in European-ancestry GWAS meta-analyses. Effects persisted after adjustment for genetic proxies of central adiposity, HbA1c, LDL cholesterol, and triglycerides, consistent with potential pathway effects on psoriatic disease, independent of metabolic effects. To determine the specificity of the observed effect, we conducted analyses of seven additional immune-mediated diseases (IMIDs). No protective effects were observed for other IMIDs, including acne and atopic dermatitis. In contrast, risk-increasing effects were observed for Crohn’s disease (OR=1.242, 95% CI 1.107-1.395, p=2.35×10^−4^) and rheumatoid arthritis (OR=1.502, 95% CI 1.350-1.672, p=1.36×10^−13^).

These findings provide genetic evidence of disease-specific, potentially direct immunomodulatory effects of GLP1R signaling on psoriasis and PsA risk, and support further mechanistic and therapeutic evaluation in randomized trials.

## Introduction

Psoriasis is a chronic, immune-mediated skin disease associated with substantial morbidity and a diminished quality of life^1^. The cumulative burden is compounded by the development of comorbidities such as psoriatic arthritis (PsA), obesity, dyslipidaemia, type 2 diabetes, and cardiovascular disease, which contribute to excess deaths in this population^2,3^. This underscores the importance of identifying treatment strategies that not only control cutaneous inflammation but also target systemic pathways relevant to the development and progression of comorbidities.

Glucagon-like peptide-1 receptor (GLP1R) agonists, originally developed for the treatment of type 2 diabetes, have demonstrated therapeutic efficacy across an expanding spectrum of cardiometabolic disorders, including obesity, cardiovascular disease, and metabolic dysfunction-associated steatohepatitis^4-6^. Notably, several conditions where GLP1R agonists have established benefits, including obesity, dyslipidaemia, fatty liver disease, obstructive sleep apnoea and type 2 diabetes, are highly prevalent in psoriasis populations^2,7^.

Beyond the central role in metabolic regulation, GLP1R signaling has also been implicated in vascular and immune biology^8^. GLP1R agonists exert pleiotropic effects, including delayed gastric emptying, appetite suppression via central nervous system pathways, enhanced insulin secretion, improved endothelial function, and reduction in Toll-like receptor– mediated inflammation^9,10^. GLP1 receptors are also expressed in multiple cell types relevant to psoriasis pathogenesis, including keratinocytes, macrophages, and endothelial cells^11^. GLP1R expression has been found to be upregulated in psoriatic lesional skin, and preclinical studies suggest that these pathways may modulate IL-17 and TNF-α signaling, key inflammatory mediators in psoriatic disease^11-13^.

Clinical studies have also reported improvements in psoriasis severity following the initiation of GLP1R agonists. A recent meta-analysis of four cohort studies and two randomized trials (n=63) found a pooled reduction in the Psoriasis Area and Severity Index (PASI) of −5.8 (95% CI: −9.5 to −2.1)^14^. An open-label, randomized study of semaglutide in patients with psoriasis and diabetes demonstrated reductions in PASI, as well as systemic markers of inflammation (IL-6 and CRP)^15^. However, existing studies are limited by small sample sizes, a lack of blinding or placebo controls, and, in some cases, entry criteria restricted to individuals already receiving systemic treatments. Taken together, these findings highlight potential clinical benefit, although it remains unclear whether any effects arise from direct immunological actions of GLP1 signaling or indirect consequences of improved metabolic control.

Mendelian randomization (MR), which uses germline genetic variation to approximate lifelong exposure to a modifiable factor, provides a complementary approach to assessing causality while mitigating confounding and reverse causation^16^. Target-focused MR with cis-expression quantitative trait loci (cis-eQTLs) genes biologically related drug target genes can provide mechanistic insights into the possible consequences of perturbing these targets in humans^17-19^. Such approaches have also been applied to evaluate the effects of genetic proxies of GLP1R expression in non-immune-mediated traits, including cancer susceptibility and progression, demonstrating their broader utility in elucidating therapeutic target biology^20^. Prior MR studies examining the effects of genetic proxies of GLP1R expression in psoriasis have reported conflicting findings, with one study showing a risk-increasing effect and another showing a protective effect, and no investigation of PsA^21,22^. These discrepancies likely stem from underpowered designs, limited case numbers, and reliance on population-based datasets with variable disease definitions, raising concerns about outcome misclassification and imprecise effect estimates.

To address these limitations, the hypothesis that GLP1 biology influences susceptibility to psoriatic disease was tested using two-sample MR applied to the most recent and largest European-ancestry GWAS meta-analyses. Firstly, the effects of genetic proxies of GLP1R expression in whole blood on psoriasis and PsA were assessed. Secondly, multivariable MR was used to determine whether associations were independent of genetic proxies of metabolic traits (including measures of central adiposity, lipids, and glycemia). Finally, to examine disease specificity, analyses were extended to seven additional immune-mediated diseases (IMIDs) involving the skin, gut, CNS, and joints.

## Results

### 1. Genetic proxies of GLP1R expression in whole blood are associated with reduced risk of cardiometabolic traits

To examine the effects of genetic proxies of GLP1R expression in whole blood on cardiometabolic traits with established real-world therapeutic benefit of GLP1R agonists, two-sample MR was performed using a genetic instrument of 22 cis-eQTLs for GLP1R expression. Genetic proxies of GLP1R expression were associated with reduced risk of type 2 diabetes (IVW OR [odds ratio] = 0.862, 95% CI = 0.840-0.885, p=4.99×10^-30^), coronary artery disease (IVW OR = 0.921, 95% CI = 0.889-0.953, p= 5.22×10^−6^), and lower levels of HbA1c (IVW β = -0.028, 95% CI = -0.037 to -0.018, p= 2.14×10^−8^), triglycerides (IVW β = -0.033, 95% CI = -0.043 to -0.024, p= 4.11×10^−11^), LDL cholesterol (IVW β = -0.035, 95% CI = -0.045 to - 0.025, p= 2.56×10^−12^), and WHRadjBMI (IVW β = -0.053, 95% CI = -0.067 to -0.039, p= 3.69×10^−14^) in GWAS meta-analysis data (Figure 2).

**Figure 1.**
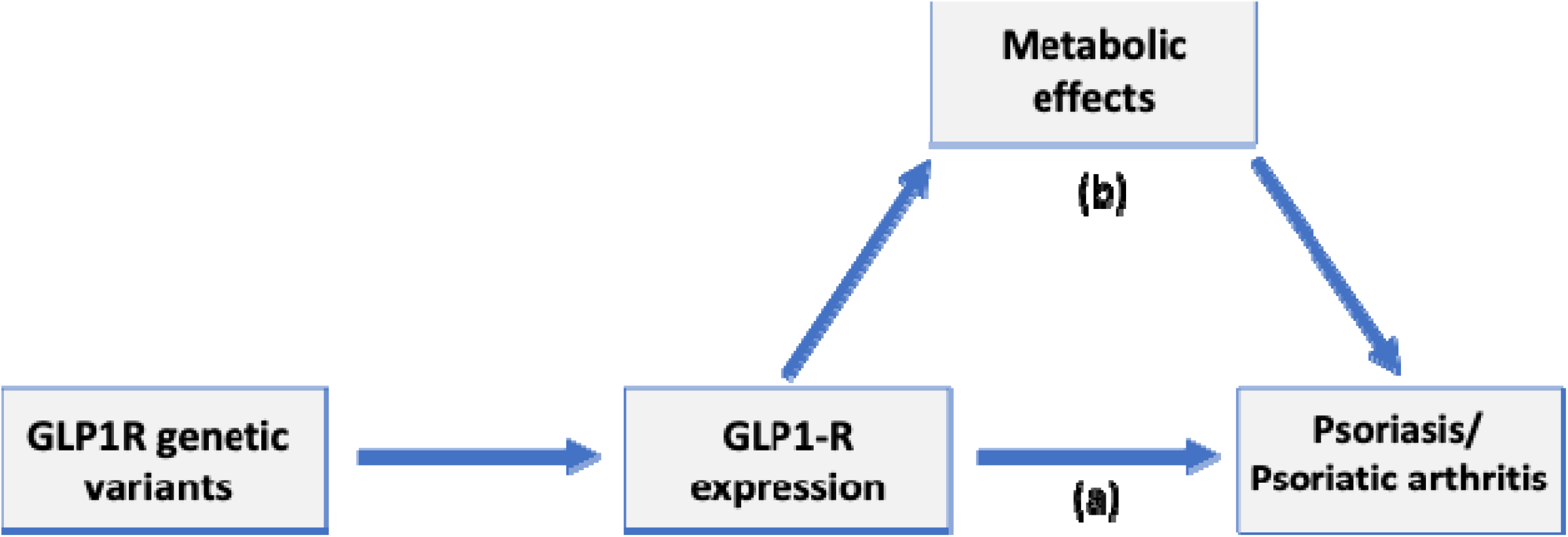
Conceptual framework evaluating the effects of genetic proxies of GLP1R expression in whole blood on psoriasis and psoriatic arthritis. (a) The use of Mendelian randomization with cis-eQTLs for GLP1R expression in whole blood as instruments to estimate effects on psoriasis and psoriatic arthritis (b) Exploring independence from indirect metabolic effects as measured by central adiposity, glycemic and lipid levels.

**Figure 2.**
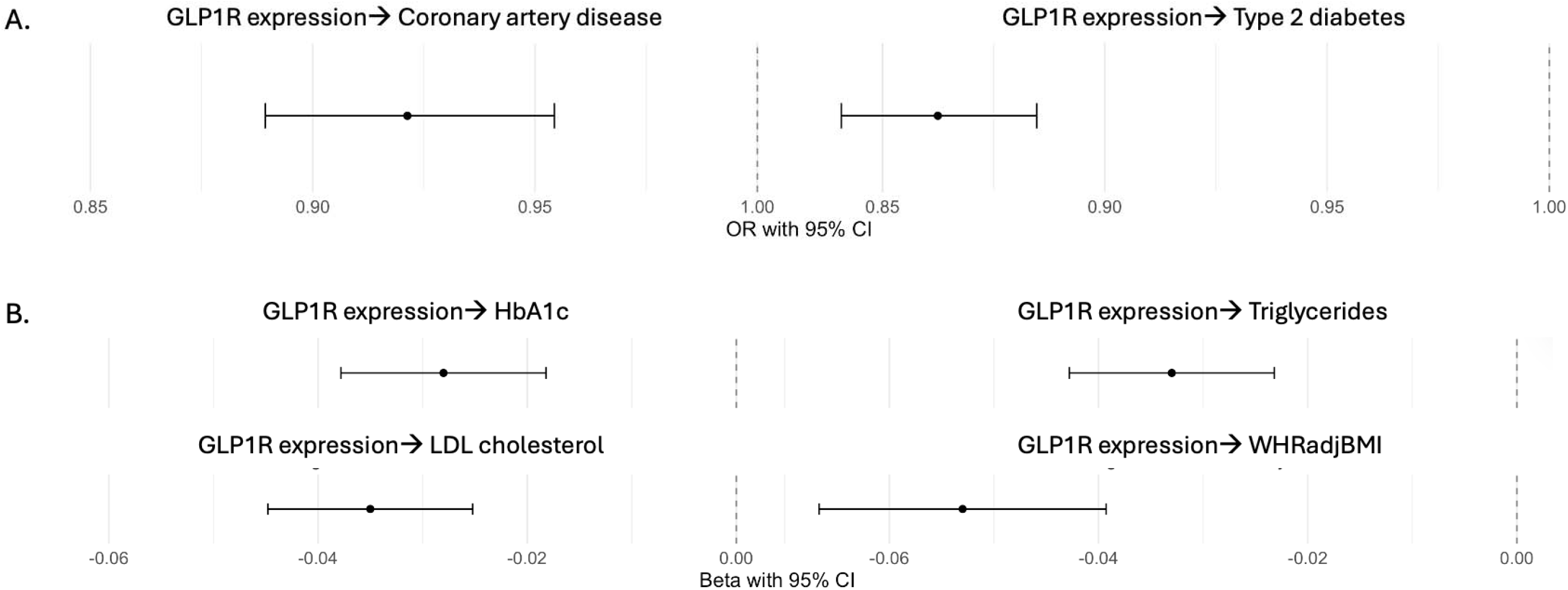
Mendelian randomization estimates for the effect of GLP1R expression in whole blood on cardiometabolic traits. Two-sample Mendelian randomization using inverse-variance weighted models was performed to estimate the effect of genetic proxies of GLP1R expression in whole blood on (a) coronary artery disease and type 2 diabetes, and (b) cardiometabolic traits, including HbA1c, triglycerides, low-density lipoprotein (LDL) cholesterol, and waist-to-hip ratio adjusted for body mass index. Effect estimates are shown as odds ratios with 95% confidence intervals for binary outcomes (a), and beta coefficients with 95% confidence intervals for continuous traits (b).

### 2. Genetic proxies of GLP1R expression in whole blood are associated with reduced risk of psoriasis and PsA, with evidence of independence from metabolic effects

Genetic proxies of GLP1R expression in whole blood were significantly associated with reduced risk of psoriasis (IVW OR = 0.723, 95% CI = 0.678-0.771, p=1.08×10^-22^) and PsA (IVW OR = 0.483, 95% CI = 0.402-0.580, p=5.33×10^-15^) in GWAS meta-analysis data (Figure 3). Findings were also consistent across different MR models used in sensitivity analysis (Appendix S1). These associations remained statistically significant following multivariable MR analyses, adjusting for genetic proxies for central adiposity (WHRadjBMI), glycaemic control (HbA1c), LDL cholesterol, and triglycerides, both individually and collectively in GWAS meta-analysis datasets (Figure 4). A modest attenuation of the effect estimate for psoriasis was observed when adjusting for all covariates simultaneously (IVW OR = 0.864, 95% CI = 0.798-0.936, p=0.003), whereas no attenuation was seen for PsA (IVW OR = 0.419, 95% CI = 0.386-0.455, p=1.43×10^-13^).

**Figure 3.**
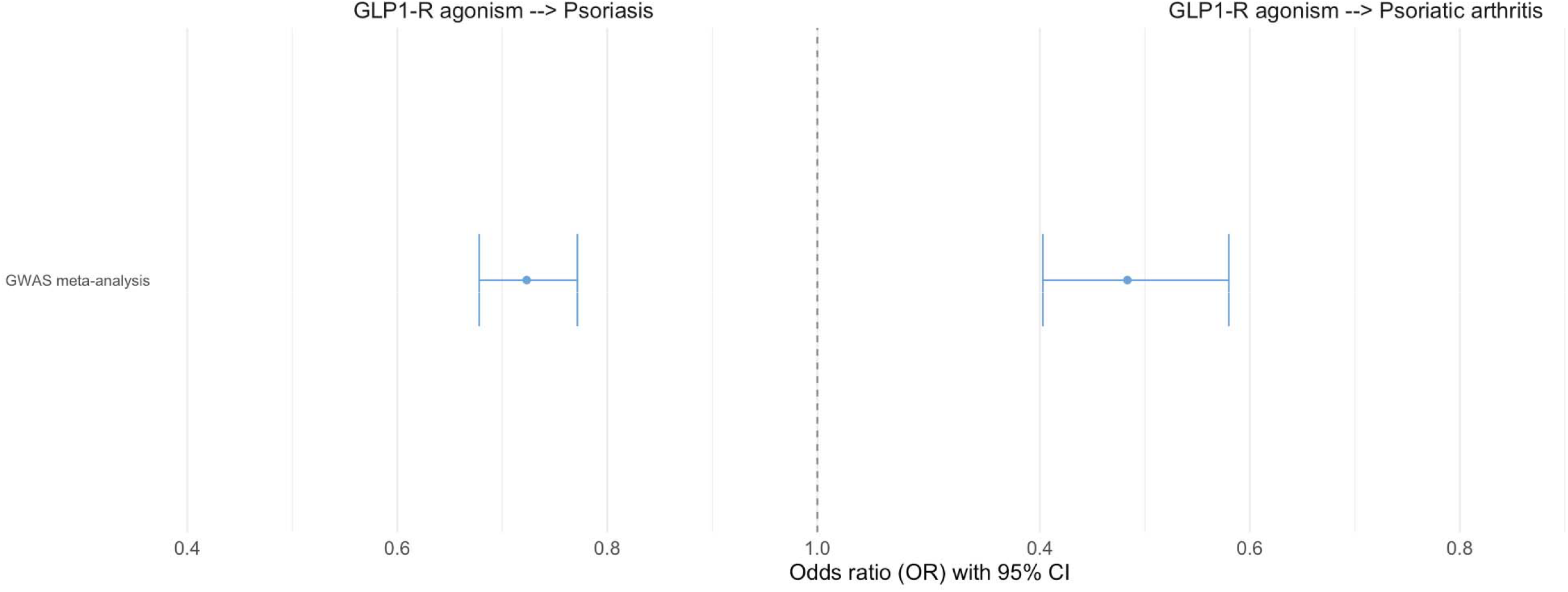
Mendelian randomization estimates for the effect of GLP1R expression in whole blood on psoriasis and psoriatic arthritis. Two-sample Mendelian randomization using inverse-variance weighted models was performed to estimate the effect of genetic proxies of GLP1R expression in whole blood on the risk of psoriasis and psoriatic arthritis in GWAS meta-analysis. Odds ratios with 95% confidence intervals are presented.

**Figure 4.**
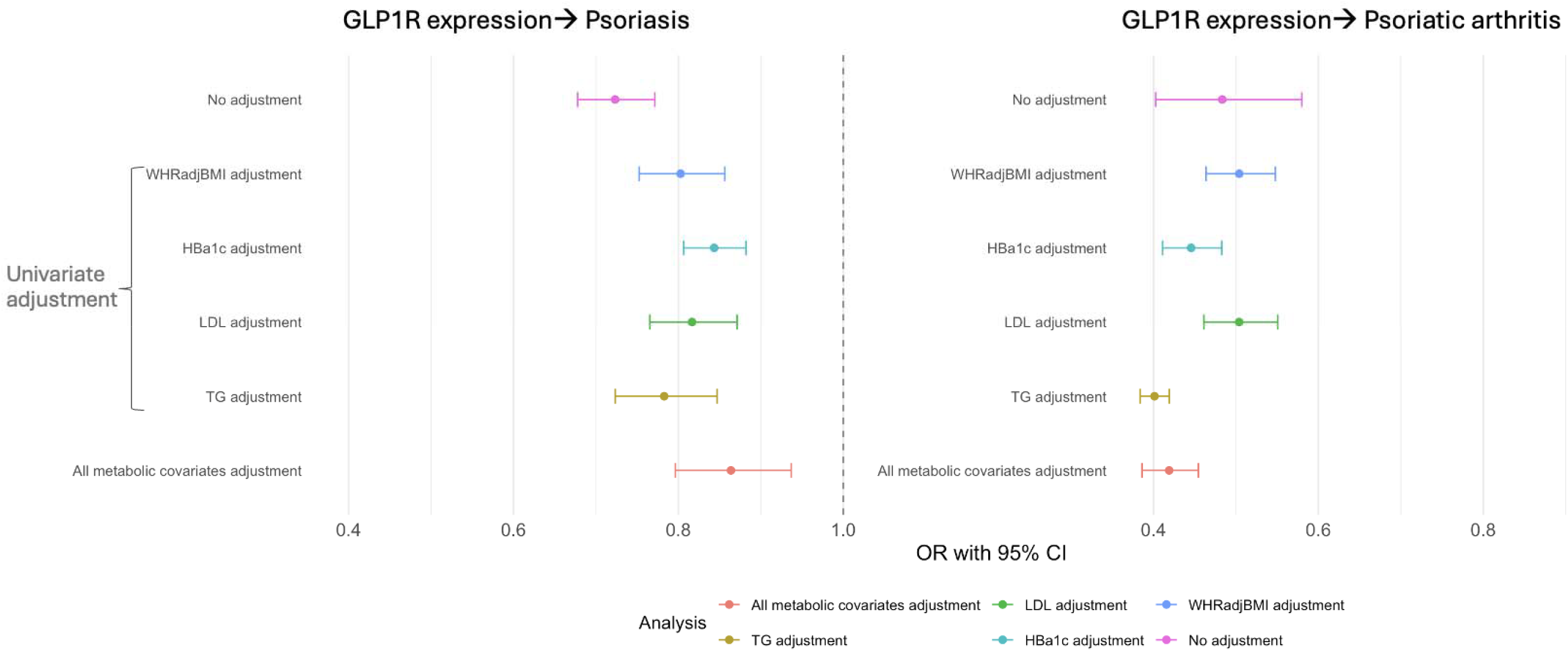
Multivariable Mendelian randomization estimates for the effect of GLP1R expression in whole blood on psoriasis and psoriatic arthritis. Two-sample Mendelian randomization (MR) using inverse-variance weighted models was used to estimate the effect of genetic proxies of GLP1R expression in whole blood on the risk of psoriasis and psoriatic arthritis in GWAS meta-analysis datasets. Odds ratios with 95% confidence intervals are shown. Models adjusting for genetic proxies of waist-to-hip ratio adjusted for body mass index (WHRadjBMI), HbA1c, low-density lipoprotein (LDL) cholesterol, triglycerides (TG) individually, and additionally all metabolic covariates collectively.

### 3. Protective effects of GLP1R expression are not observed across other immune-mediated diseases

No protective effects of genetic proxies of GLP1R expression were observed for seven other common IMIDs in GWAS meta-analyses. No effects were observed on other skin or joint diseases, in contrast to the findings for psoriasis and PsA, nor was there evidence of a protective effect on inflammatory bowel disease or multiple sclerosis risk (Table 1). Conversely, genetic proxies of GLP1R expression were associated with increased risk of Crohn’s disease (IVW OR = 1.242, 95% CI = 1.107-1.396, p=2.35×10^-04^) and RA (IVW OR = 1.502, 95% CI = 1.350-1.672, p=1.36×10^-13^) (Table 1, Appendix S1).

**Table 1.** Mendelian randomization (MR) analysis of GLP1R expression in whole blood and risk of immune-mediated diseases. Odds ratios (ORs) and 95% confidence intervals (CIs) represent the estimated effect of genetic proxies of GLP1R expression in whole blood on the risk of seven immune-mediated diseases using MR in GWAS meta-analysis datasets. Inverse-variance weighted ORs are shown, and p-values. Associations that remain statistically significant after Bonferroni correction for multiple testing (p < 0.0056) are shown in bold and denoted by an asterisk (*).

| Outcome (immune-mediated disease) | OR (95% CI) | P-value |
| --- | --- | --- |
| Acne | 1.04 (0.96–1.13) | 0.379 |
| Asthma | 1.03 (0.99–1.07) | 0.187 |
| Atopic dermatitis | 1.01 (0.96–1.05) | 0.835 |
| Crohn's disease* | 1.24 (1.11–1.40) | **2.35x10<sup>-4</sup>** |
| Multiple sclerosis | 0.89 (0.80–1.00) | 0.056 |
| Rheumatoid arthritis* | 1.50 (1.35–1.67) | **1.36x10<sup>-13</sup>** |
| Ulcerative colitis | 1.01 (0.96–1.05) | 0.835 |

## Discussion

This study provides genetic evidence that GLP1R pathway activity influences immune disease susceptibility in a disease-specific manner, being associated with lower risk of psoriasis and PsA, no effect on acne and atopic dermatitis or multiple sclerosis, and higher risk of Crohn’s disease and RA. Persistence of associations with psoriatic disease after multivariable adjustment for central adiposity, glycaemia, and lipids suggests that the effects may not be fully explained by metabolic pathways, raising the possibility that GLP1R signaling may also influence psoriatic disease biology through direct immunomodulatory mechanisms. These genetically anchored findings are hypothesis-generating and may predict the effects of long-term modulation of the GLP1R pathway but require triangulation with other forms of evidence and confirmation in randomized clinical trials.

The protective effects on psoriasis reinforce emerging clinical and mechanistic evidence^7,13^. A small RCT of liraglutide in patients with both type 2 diabetes and psoriasis demonstrated improvement in disease severity^23^. These findings support further investigation of GLP1R agonists as potential immunomodulatory therapies in individuals with psoriasis. Given the high burden of cardiovascular and metabolic comorbidities in these populations^24,25^, dual benefit on both inflammatory and metabolic pathways would represent a clinically valuable therapeutic profile.

Effects of GLP1R expression on psoriasis were modestly attenuated after adjustment for genetic proxies for BMI, HbA1c, LDL cholesterol, triglycerides, and when all covariates were included together in a single model, but associations remained statistically significant. In contrast, the protective effect on PsA was not attenuated by adjustment for any of these traits. However, these findings should be interpreted with caution, as the genetic instruments used for metabolic traits are only partial predictors of the corresponding phenotypes, and incomplete adjustment could allow residual confounding. These findings may suggest a pleiotropic role of GLP1R signaling beyond metabolic perturbation^9^. Evaluation of immune tissue compartments such as skin and synovium will be important to help elucidate the mechanisms underlying these associations.

The risk-increasing associations for Crohn’s disease and RA emphasize potential heterogeneity of immune mechanisms across diseases and caution against assuming uniform benefit of pathway perturbation. The risk-increasing effects in Crohn’s disease contrast with some real-world studies, which have shown no worsening of disease activity in individuals with IBD treated with GLP1R agonists^26-28^. Similarly, established targeted biologic therapies such as interleukin-17 (IL-17) pathway inhibitors have been associated with an increased risk or exacerbation of Crohn’s disease, underscoring the context-dependent effects of immune pathway modulation^29^. The apparent discrepancy may reflect differences in the underlying mechanisms influencing disease onset versus disease control. Notably, both high and low body mass index are associated with Crohn’s disease, and metabolic shifts may alter risk in susceptible individuals^30^. Furthermore, while a case report has described the development of RA following treatment with a GLP1R agonist, existing data remain sparse^31^. The opposing effects observed between PsA and RA highlight the heterogeneity in immune mechanisms across IMIDs and suggest differential effects of GLP1R modulation depending on disease context. Given these findings, it is important to maintain vigilance for potential effects on Crohn’s disease and RA in pharmacovigilance and epidemiological studies, in order to validate potential adverse signals and ensure the safe use of GLP1 pathway-modulating therapies in individuals with, or at risk of, these conditions.

Limitations include that MR estimates reflect lifelong genetic perturbation rather than the temporal dynamics of relatively shorter-term pharmacological treatment^32^. Analyses were also restricted to individuals of European ancestry, so generalisability to other ancestral groups requires evaluation. Additionally, the GLP1R expression instrument is based on whole-blood cis-eQTLs and may not fully capture tissue-specific regulation in skin, synovium, brain, or gut. It should also be recognized that using a cis-eQTL for GLP1R expression as a proxy for pathway activity simplifies the underlying biology, as it reflects a lifelong genetic tendency toward altered receptor expression rather than the dynamic, feedback-regulated nature of GLP1 signaling. Nonetheless, such instruments can provide useful insight into the predominant direction and potential consequences of pathway perturbation at the population level. Nevertheless, GLP1 is a circulating hormone with established vascular and inflammatory relevance, and circulating GLP1 levels correlate with cardiometabolic and endothelial phenotypes, supporting the translational relevance of a blood-based proxy for pathway activity^33^. Finally, MR provides average population-level effects and cannot exclude effect heterogeneity by metabolic status, treatment history, or background disease.

Strengths of this study include a target-focused instrument with strong biological plausibility, validation against cardiometabolic positive controls, large and well-powered GWAS meta-analyses for psoriasis and PsA, the specific protective effect observed in particularly in psoriasis (and not other IMIDs), and robustness across multiple sensitivity estimators. Together, these features strengthen causal inference and enhance the translational relevance of the findings.

In summary, this study’s findings position GLP1 signaling as a biologically plausible and potentially modifiable pathway in psoriatic disease. These results provide a mechanistic rationale to test durable GLP1 pathway modulation in the treatment of psoriatic disease. Future work should resolve tissue and cell type mechanisms, evaluate generalizability across ancestries, and define subgroups most likely to benefit. Collectively, these data extend GLP1 biology beyond metabolism, identifying an immune-relevant pathway with testable therapeutic potential in psoriatic disease.

## Methods

This method’s description follows the Strengthening the Reporting of Observational Studies in Epidemiology Using Mendelian Randomization (STROBE-MR) guidelines^34^.

### Study design

This investigation uses MR to explore the association between genetic proxies of GLP1R expression in whole blood on psoriatic disease risk.

MR is a statistical method used to assess causality and make inferences about the potential causal relationships between an exposure (or risk factor) and an outcome of interest^35^. MR leverages genetic variants, typically single-nucleotide polymorphisms (SNPs), as instrumental variables. This approach circumvents common limitations inherent in observational studies, such as unmeasured confounding and reverse causation, because genetic alleles are randomly allocated at conception and remain fixed throughout an individual’s life, analogous to a natural RCT. These inherent features of MR provide a strong foundation for estimating causal effects.

The primary analytical framework for this study involves two-sample MR. In this design, associations between genetic variants and the exposure of interest (in this case GLP1R expression in whole blood) are estimated from one dataset, while associations between genetic variants for GLP1R expression and the outcome (e.g., psoriasis) are derived from a second, distinct dataset (or separate meta-analysis of datasets). To mitigate potential biases arising from population stratification, all analyses within this study were restricted to individuals of European ancestry. The key study steps are listed below (Figure 1):

1. **Positive control analyses -** To ensure the effectiveness and biological fidelity of the selected genetic instruments, a positive control analysis was performed. This involved examining the association between the genetic proxies of GLP1R expression and six cardiometabolic traits with known therapeutic benefits in clinical trials and real-world practice.
2. **Primary analyses-** The association between genetic proxies of GLP1R expression and risk of psoriasis and PsA was examined using two-sample MR in large-scale GWAS meta-analysis data.
3. **Multivariable MR analyses-** For psoriasis and PsA, multivariable MR was performed to assess whether observed associations were independent of key metabolic traits influenced by GLP1R pathway activity in GWAS meta-analysis datasets. Genetic proxies for HbA1c, triglycerides, and LDL cholesterol, and waist-to-hip ratio adjusted for BMI (WHRadjBMI) were adjusted for given the known strong association between central adiposity and psoriasis^36^.
4. **Exploratory analyses of other IMIDs-** To assess disease specificity, the effects of genetic proxies of GLP1R expression were examined on seven additional common immune-mediated diseases affecting the skin, gut, CNS, and joints.

### Data sources, genetic instruments and study populations

The exposure was genetic proxies of GLP1R expression in blood using 22 independent cis-eQTLs from eQTLGen (linkage disequilibrium r^2^ < 0.001) associated with GLP1R expression in whole blood (p<5×10^−8^) from the eQTLGen consortium (n=31,684, Appendix S1)^37^.

GWAS summary statistics were obtained from publicly available large-scale consortia. For each trait, summary-level data from the most recent and well-powered GWAS meta-analyses of European ancestry were used. The primary cardiometabolic outcomes assessed were type 2 diabetes (62,892 cases, 596,424 controls)^38^, coronary artery disease (181,249 cases; 1,165,690 controls)^39^, HbA1c (n=144,060)^40^, triglycerides (n= 1,320,016)^41^, low-density lipoprotein (LDL) cholesterol (n= 1,320,016)^41^, and waist-to-hip ratio adjusted for BMI (WHRadjBMI) (n=694,649)^42^. Nine IMIDs were evaluated: acne (20,165 cases; 595,231 controls)^43^, asthma (19,954 cases; 107,715 controls)^44^, atopic dermatitis (60,653 cases; 804,329 controls)^45^, Crohn’s disease (40,266 cases; 28,072 controls)^46^, multiple sclerosis (47,429 cases; 68,374 controls)^47^, psoriasis (36,466 cases; 458,078 controls)^48^, PsA (cases: 5,065, controls: 21,286)^49^, rheumatoid arthritis (RA) (29,880 cases; 73,758 controls)^50^ and ulcerative colitis (33,609 cases; 45,975 controls)^46^.

### Assumptions

A genetic variant can be considered as an instrumental variable for a given exposure if it satisfies the instrumental variable assumptions: 1) it is associated with the exposure, 2) it is not associated with the outcome through confounding pathways, and 3) it does not affect the outcome except potentially via the exposure.

### Statistical analysis

Data harmonization was carried out to ensure allele correspondence between the exposure and the outcome for two-sample MR analyses.

The primary statistical method for evaluating MR associations was the Inverse-variance weighted (IVW) MR. Estimates for binary outcomes were reported as odds ratios (ORs) per standard deviation increase in GLP1R expression with corresponding 95% confidence intervals (CIs). For continuous outcomes, beta coefficients and 95% CIs were reported. A Bonferroni-corrected significance threshold of p < 0.0056 was used to account for multiple testing across nine IMID outcomes (including primary endpoints of psoriasis and PsA).

For psoriasis and PsA, additional multivariable MR (MVMR) analyses were performed to assess whether the observed associations were independent of metabolic traits commonly influenced by GLP1 pathway activity in GWAS meta-analysis datasets. Specifically, genetic proxies for WHRadjBMI, HbA1c, triglycerides, and LDL cholesterol were included as covariates in both individual and joint models. This approach allowed assessment of whether the observed associations with psoriasis and PsA represent direct effects of GLP1R expression, independent of genetic proxies for metabolic traits.

### Assessment of assumptions

The F-statistic for GLP1R expression used in two-sample MR was 53.0. All genetic instruments used had an F-statistic of ≥ 10, exceeding the conventional threshold for sufficient instrument strength and supporting the validity of the relevance assumption of MR^51^.

### Sensitivity analyses

Results have potential for bias if instrumental variables exhibit horizontal pleiotropy, influencing the outcome through causal pathways other than the exposure, thus violating the instrumental variable assumptions as above^52^. Additional MR analyses were therefore performed using methods with differing assumptions about instrument validity. These included the simple and weighted median methods (robust when up to 50% of instruments are invalid), MR-Egger (which accounts for directional pleiotropy), and robust approaches such as penalised MR-Egger, robust IVW, and MR-Lasso. Consistency in the direction and magnitude of effects across these methods was used to support the validity of the primary IVW estimates.

### Study approval

The primary analysis in this study utilised publicly available summary-level data from previously published GWAS. As these analyses did not involve individual participant data or direct contact with human subjects, this research was exempt from institutional ethical review.

### Software

All MR analyses were performed using R (version 4.1.2) (R Foundation for Statistical Computing, Vienna, Austria) using the “TwoSampleMR”, “MendelianRandomization” and “MVMR” packages^53,54^.

## Data availability

Two-sample MR was performed using publicly available summary statistics that can be downloaded from the GWAS catalogue (https://www.ebi.ac.uk/gwas/) and/or can be downloaded from the GWAS catalogue obtained from the corresponding author of the GWAS on reasonable request. All data supporting the findings of this study are presented within the manuscript and the supplementary file. Additional details and code used for Mendelian randomization analyses are available from the corresponding author upon reasonable request.

